# Heightened non-testing risk mitigation blood donor screening with retrospective transcription-mediated amplification testing during a 2023 local Florida malaria cluster

**DOI:** 10.1101/2025.02.21.25322106

**Authors:** David J. Sullivan, Marion Lanteri, Vanessa Bres, Measa Hanhan, Marlene Pagan, Alejandro Rey, Korena Thomas, Betsy Sasnett, Rita A. Reik

## Abstract

**Background:** Transfusion-transmitted malaria risk in the USA is estimated at less than one per ten million blood donations or about 1 case every 2 years. A total of 18 autochthonous malaria infections have been reported in the USA since 2003: eight cases in 2003 and ten cases in 2023 with fifteen reported from Florida. From May to July 2023 in Sarasota County, Florida, symptomatic *P. vivax* malaria was detected in seven individuals with no recent malaria travel history.

**Study Design and Methods:** In the absence of an FDA-approved blood donor screening test for malaria at the time, the local blood center responded to this 2023 cluster by implementing a non-testing risk mitigation strategy of escalating blood donor screening measures and pathogen reduction. Potential blood supply safety enhancement by use of additional nucleic acid testing (NAT) on samples from donors in impacted areas was studied retrospectively using the Procleix Plasmodium Assay, a transcription-mediated amplification detecting five human *Plasmodium* species 18S ribosomal RNA with a limit of detection ranging from 2-7 infected erythrocytes per mL.

**Results:** Among the 435 valid study samples from accepted healthy blood donors, screened with heightened non-testing measures in the 4-6 mile radius of malaria-impacted area, all samples were nucleic acid test non-reactive.

**Discussion:** The blood center strategy in response to the local 2023 malaria outbreak maintained the blood supply safety with no transfusion-transmitted malaria and no retrospective evidence of *Plasmodium* in the 435 valid samples collected from blood donors residing in impacted areas.

## Introduction

Blood donors represent a population of healthy individuals comprising about 6.5 million people donating an estimated 11.8 million units of whole blood and red cell products annually in the U.S^1^. Presently, the Uniform Donor History Questionnaire (UDHQ) and an in-person mini-physical are used to screen prospective blood donors. Additionally, U.S. blood donors are deferred within 3 months of travel to a malaria endemic area. Former malaria endemic residents and recent malaria cases are deferred for 3 years. Using these measures, there have been only 13 transfusion-transmitted malaria (TTM) cases between 2000 and 2021, (average 0.59/year) or less than 1 in 10 million blood donations in the U.S.^2,3^.

Malaria elimination in the U.S. in the 1950s had little influence on Anopheles species mosquito populations over the past 70 years^4^. Although *Anopheles* mosquitoes are widespread in the U.S, most malaria cases are travel-associated^3^. The presence of the mosquito vector in the U.S. has been associated with small self-contained rare local malaria outbreaks recently in 2003 in Palm Beach County Florida with eight *P. vivax* cases^5^, then twenty years later in 2023. Seven of the ten 2023 malaria cases were *P. vivax* cases from Sarasota county Florida^6,7^. A single case of *P. falciparum* was reported in Maryland^8^ with two more P. vivax cases in Texas^7^ and Arkansas^9^.

Local malaria is geographically restricted as most mosquitoes reside within 1-2 kilometers of larval hatching for the 2–6-week normal total lifespan of the female mosquito. In the Sarasota Florida 2023 cluster, all of the 7 local malaria *P. vivax* cases remained within a 4-mile radius. There were three homeless individuals among the 7 without history of travel. After notification by the Florida Department of Health of local malaria confined to a specific location, the blood center (BC) implemented its “Vector-borne Pathogen Mitigation Strategy” (VPMS) which consists of heightened donor education and screening measures, product embargoes and donor call backs, along with pathogen reduction of eligible products (INTERCEPT^™^ for Platelets and Plasma, Cerus corporation, Concord, USA) for blood donors in the affected area. The VPMS has been successfully employed by the blood center since 2013 during outbreaks of dengue, chikungunya, Zika and malaria with modifications based on known characteristics of the vector and pathogen involved. Additionally, blood samples from 435 donors residing in the impacted area from July 2023 to October 2023 were retained for future investigation aimed at identifying subclinical infections using a sensitive *Plasmodium* nucleic acid test (NAT).

## Methods

### Retrospective testing of collected samples

The donor study protocol was approved by the Institutional Review Board Advarra with protocol number Pro00079961. Donors were initially screened and accepted using the non-testing VPMS (Table 1). The collections during that period ranged from 1 to 21 donors a day on 77 days of the 91-day span over the three months. Whole blood samples from 436 blood donors residing in the impacted area were harvested and individual whole blood lysates were manually prepared by mixing 0.9 mL of fresh whole blood with 2.7 mL of Parasite Transport Media (PTM), a proprietary lysis and RNA-stabilizing solution (Grifols Diagnostic Solutions Inc), in an individual donor lysate (IDL) tube. IDLs were then frozen at ≤-20°C until testing was performed.

**Table 1.**
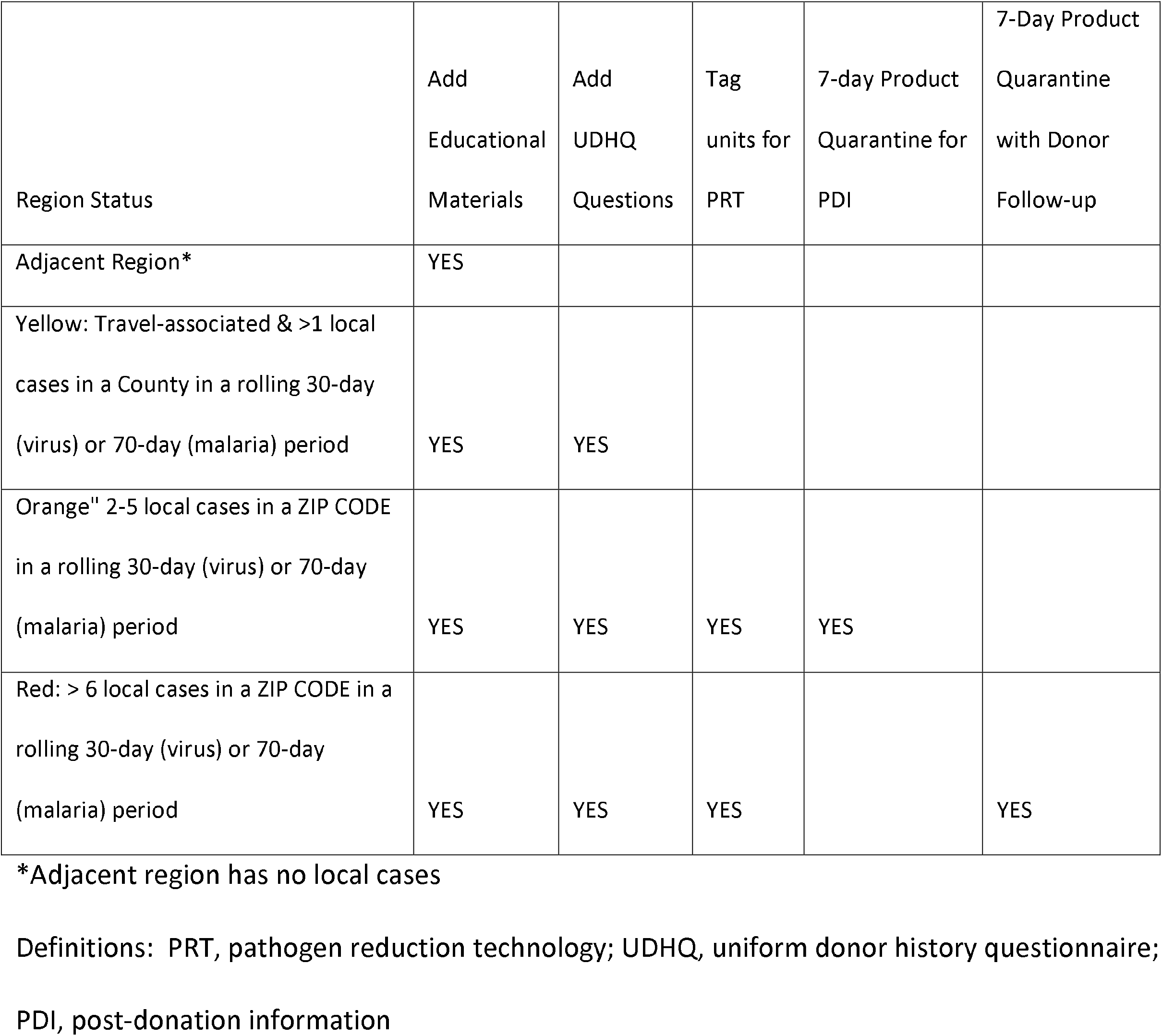
FL BC “Vector-borne Pathogen Mitigation Strategy” including Malaria.

### Sample Testing

Samples were retrospectively tested for *Plasmodium* using a highly sensitive nucleic acid test (Procleix Plasmodium Assay, Grifols Diagnostic Solutions Inc, San Diego, USA).

The Procleix Plasmodium Assay is a qualitative *in vitro* transcription-mediated nucleic acid amplification (TMA) test for the detection of 18S ribosomal RNA (rRNA) from five *Plasmodium* species (*P. falciparum, P. ovale, P. vivax, P. malariae*, and *P. knowlesi*). This assay is not yet available for blood donor screening in the U.S. but used in countries/regions recognizing the CE mark. The assay is conducted on a fully automated Procleix Panther System (Grifols Diagnostic Solutions Inc, San Diego, CA). Frozen lysates were thawed and loaded on the Procleix Panther System, which uses 0.5 mL of lysate to perform the Procleix Plasmodium Assay. This involves three main steps: magnetic-based target capture, RNA target amplification by transcription-mediated amplification, and chemiluminescent detection. The assay specific software determines if the sample is reactive or nonreactive for the presence of *Plasmodium* RNA. Specimens with an Analyte Signal/ Cutoff (S/CO) ≥1.00 are considered reactive^10^.

## Results

### Local malaria outbreak in Sarasota Florida 2023

An imported malaria case with disease onset in April was diagnosed in 2023^7^. A second P. vivax local malaria case (homeless status unknown) presented in May with a 14-day history of symptoms. After two weeks with no malaria cases, 4 more local cases presented in the middle of June with two more local cases late June and early July (**Figure 1**). The prepatent (time of symptomatic parasitemia from infected mosquito bite) is approximately 12 to 14 days in *P. vivax* case presentations^11,12^. In order to transmit to humans, the mosquito needs to acquire the P. vivax infectious gametocytes in a previous bloodmeal at least 8 days prior to transmission bite, because of a temperature dependent development process and relocation from midgut to salivary glands in the mosquito^13^. The average daily temperature in April was 25⍰C, in May 26⍰C, in June 29⍰C and July 31⍰C. Because infected mosquitoes can live from 1 to 7 weeks, all the local cases can be explained by feeding on the second *P. vivax* case who has a 14-day symptom period before heath evaluation. In a genome sequence analysis by the University of South Florida working with the Florida Health Department, malaria cases 4, 6, 7 and 8 who presented as the last four cases were identical or clonal^14^. The initial *P. vivax* imported case did not have evaluable DNA material. If the imported malaria case 1 had been parasitemic and asymptomatic for 2 weeks prior to symptom onset then all the cases were related. Another more remote possibility is the second case acquired the *P. vivax* from an asymptomatic immigrant/traveler. The CDC investigated 30 people with syndromes suggestive of malaria, but all were diagnosis test negative with the BinaxNOW point of care rapid diagnostic tests which have a sensitivity of 10,000 to 100,000 per mL of blood^7^. In the local area of 407 mosquitoes tested, 3 were positive for *P. vivax* DNA in mosquito in midguts, but all were negative for infected sporozoites stages^7^.

**Figure 1.**
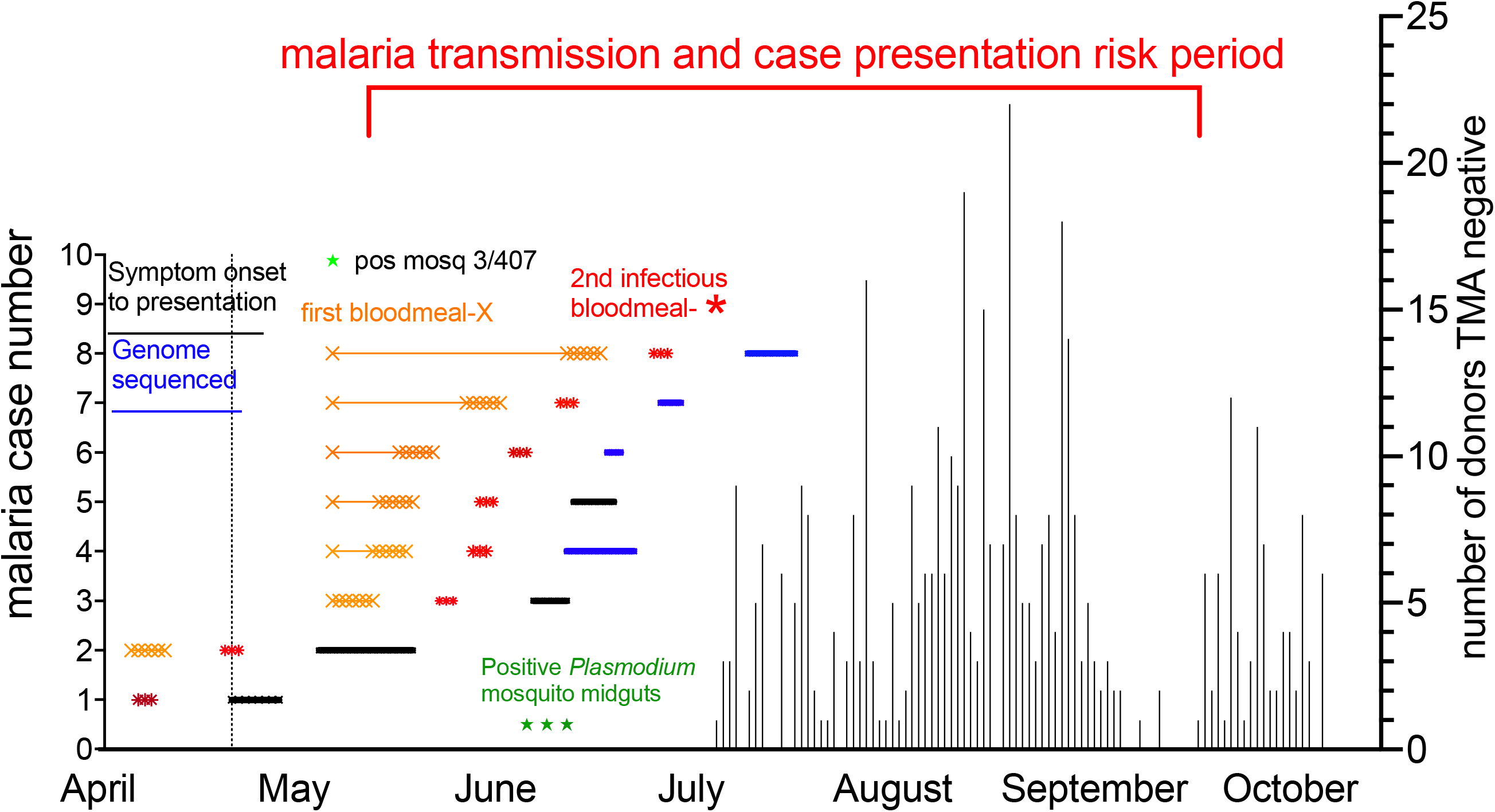
Timeline of *P. vivax* cases, range of mosquito feeding for acquisition and transmission and risk period for other malaria cases. Malaria case 1 (April presentation) was infected by a mosquito at least a week prior to case presentation and was not sequenced. Case 2 (May presentation) with 14 days of previous symptoms during which this person could have infected other mosquitoes. The gold X represent the range if initial mosquito acquisition of *P. vivax* with subsequent minimum blood feeding transmission a week before clinical case presentation represented by red *. In the local area of 407 mosquitoes tested 3 were positive for *P. vivax* DNA in mosquito in midguts, but negative for infected sporozoites stages. Represented buy green stars in early June. The right Y-axis noted the number of *Plasmodium* test negative donors over 3 months indication no measurable contamination of blood supply. The time period of maximum risk is June to September for blood supply.

### Donor malaria testing

The 436 donors from Sarasota had blood collected from July 2023 to October 2023. They were outside deferral windows as well as asymptomatic (i.e. not febrile) and passed heightened blood donor screening per the BCVPMS, and their eligible products were pathogen-reduced as indicated and released into the blood supply. The Procleix Plasmodium Assay analyzes 500 µL of whole blood lysate for the detection of 18s ribosomal RNA, which would be present in thousands of copies per cell, if one infected red blood cell was captured in the whole blood lysate. Out of 436 samples, five samples had an initial invalid result, of which four samples were valid upon re-test and one was invalid. The overall final validity was 99.77% (435/436). All 435 samples with a valid results collected in the local area from July 2023 to October 2023 were all non-reactive for *Plasmodium* (**Figure 1**).

## Discussion

In reviewing the tempo and location of the local cases observed in the 2023 FL malaria cluster, we posit that the second local case infected subsequent mosquitoes and initiated the genetically identical infections occurring in people up to the first week of July from mosquitoes feeding two months previously. In this case the risk period for malaria in the blood supply could be predicted to primarily extend into September. This pattern proved to be consistent with the known tendency for U.S. malaria clusters to remain small and local.

Also of note is that although a total of 18 local (autochthonous) malaria cases (17 due to *P. vivax* and 1 due to *P. falciparum*) transmissions have been recorded in the U.S. since 2003^3,15^, during the past 20 years there have been only 13 cases of TTM with 9 *P. falciparum*, 2 *P. malariae* and 2 *P. ovale* with no *P. vivax*^16,17^. Additionally, all traced donors were former malaria endemic residents, and none were U.S. citizens traveling back from malaria endemic areas^17^. This contrasts with the 93 TTM cases from 1963 to 1999 where 33 were *P. falciparum*, 25 *P. vivax*, 25 *P. malariae* and 5 *P. ovale*^18^. In this group the longest time from travel to a malaria region and donation of malaria infected blood was 44 years in the case of *P. malariae*, 5 years in the case of *P. falciparum*, 7 years in the case of *P. ovale* and 2.5 years in a case of *P. vivax* with most outside of the deferral interval of 3 years^18^. In that last century time period only 3 were U.S. citizens returning from travel and subsequently becoming infected blood donors. Ten of the 93 TTM recipients died from severe malaria^18^.

Preparedness for emerging infectious disease outbreaks is one of the strategic initiatives undertaken by the BC to maintain the safety and availability of the blood supply during local vector-borne agent outbreak. Indeed, the blood center is mostly collecting blood in FL, a U.S. state known to be impacted by local outbreak of mosquito-borne agents. Past FL experience includes responses to outbreaks of dengue, chikungunya and Zika viruses as well as malaria. Close collaboration with the local health department and CDC has allowed for appropriate communication for optimum reactivity to local emerging threats. As vector-borne outbreaks are unpredictable, BC developed the VPMS which was used successfully several times over the past twelve years including deployment during the 2023 malaria cluster. Although the VPMS, has proven effective to date, with no cases of transfusion-transmission associated with its use during outbreaks in FL, concerns remain regarding subclinical blood donor infections. Similarly, donor follow up after donation only partially addresses the risk associated with donations collected during the pre-symptomatic phase of infection due to variability in symptom development times.

Another mitigation strategy employed by the BC during clusters and outbreaks is the use of an approved amotosalen and ultraviolet light pathogen reduction (PR) system for the inactivation of infectious agents in platelets and plasma^19^. Such a system has been in use at the BC since 2016. A different nonapproved PR with amustaline and glutathione adequately inactivates erythrocytic *Plasmodium* among other species^20^. Over the period July to October platelets and plasma were treated with PR maintaining the safety of transfused patients.

With the implementation of the VPMS and the containment efforts of the Florida health department, the blood supply was not significantly impacted by the local outbreak, as demonstrated by highly sensitive NAT testing non-reactive results in the 435 valid donor samples from the area and no TTM. From a blood center perspective, the highly sensitive Procleix Plasmodium Assay offers the opportunity to be utilized to enhance blood safety and during local malaria outbreaks.

## Data Availability

All data produced in the present study are available upon reasonable request to the authors

## Notes

Source of support OneBlood, Inc., Grifols Diagnostic Solutions Inc., Creative Testing Solutions, Bloomberg Philanthropies

### Competing Interest Statement

DJS is Founder, Board Member, and stock/option owner of AliquantumRx(macrolide for antimicrobial and malaria use); coinventor on USP 7,270,948 (Detection of malaria parasites by laser desorption mass spectrometry), USP9,568,471 (Malaria diagnosis in urine), USP 9,642,865 (New angiogenesis inhibitors), and PCT/US2015/046665 (Salts and polymorphs of cethromycin for the treatment of disease); and has received royalties from Binax Inc/D/B/A Inverness Medical for plasmids for HRP2 and aldolase for malaria diagnostic kit. ML and AR are employees of CTS; VB and MH are employees of Grifols Diagnostic Solutions Inc; RAR is an employee of OneBlood and serves on the Medical Advisory Board of Creative Testing Solutions; MP, KT, BS are employees of OneBlood; OneBlood is part owner of, and has representation on the Board of Directors of Creative Testing Solutions

### Funding Statement

OneBlood, Inc., Grifols Diagnostic Solutions Inc., Creative Testing Solutions, Bloomberg Philanthropies

### Author Declarations

The donor study protocol was approved by the Institutional Review Board Advarra with protocol number Pro00079961.

